# Enhancing Disease Risk Gene Discovery by Integrating Transcription Factor-Linked Trans-located Variants into Transcriptome-Wide Association Analyses

**DOI:** 10.1101/2023.10.10.23295443

**Authors:** Jingni He, Deshan Perera, Wanqing Wen, Jie Ping, Qing Li, Linshuoshuo Lyu, Zhishan Chen, Xiang Shu, Jirong Long, Qiuyin Cai, Xiao-Ou Shu, Wei Zheng, Quan Long, Xingyi Guo

## Abstract

Transcriptome-wide association studies (TWAS) have been successful in identifying disease susceptibility genes by integrating cis-variants predicted gene expression with genome-wide association studies (GWAS) data. However, trans-located variants for predicting gene expression remain largely unexplored. Here, we introduce transTF-TWAS, which incorporates transcription factor (TF)-linked trans-located variants to enhance model building. Using data from the Genotype-Tissue Expression project, we predict gene expression and alternative splicing and applied these models to large GWAS datasets for breast, prostate, and lung cancers. We demonstrate that transTF-TWAS outperforms other existing TWAS approaches in both constructing gene prediction models and identifying disease-associated genes, as evidenced by simulations and real data analysis. Our transTF-TWAS approach significantly contributes to the discovery of disease risk genes. Findings from this study have shed new light on several genetically driven key regulators and their associated regulatory networks underlying disease susceptibility.

## Introduction

Approximately 90% of risk variants identified in genome-wide association studies (GWAS) are located in noncoding or intergenic regions, which suggests that they may affect cancer risk by dysregulating gene expression (1–10). Fine-mapping of genetic risk loci, along with functional experiments provide strong evidence that regulatory variants in linkage disequilibrium (LD) with GWAS-identified risk variants disrupt DNA binding affinities of specific transcription factors (TFs) and modulate expression of susceptibility genes (11–22). Thus, identifying TFs, whose DNA bindings are altered by risk-associated genetic variations, and their controlling genes can greatly improve the understanding of transcriptional dysregulation in human diseases and cancers (23–26). A pioneering study analyzed Chromatin immunoprecipitation followed by sequencing (ChIP-seq) data for TFs such as FOXA1 in multiple breast cancer cell lines to investigate the binding of GWAS-identified risk variants (21). The study suggested that regulatory variants confer to breast cancer risk by mediating their altered binding affinities. Subsequent studies have revealed multiple breast cancer risk-associated TFs such as ESR1, MYC, and KLF4 (20,27) through interrogating data on gene expression, TF ChIP-seq, and GWAS-identified risk variants. We have recently conducted a comprehensive analysis of TF ChIP-seq and GWAS data for breast cancer, and developed an analytical framework to identify TFs that contribute to breast cancer risk. Our study revealed that the genetic variations of 22 TFs were significantly associated with breast cancer risk and highlighted genetic variations of TF-DNA bindings (particularly for FOXA1) underlying breast cancer susceptibility (28).

Transcriptome-wide association studies (TWAS) have successfully uncovered large numbers of putative susceptibility genes for cancers and other diseases, and many of these genes have been further supported by functional experiments (29–33). In TWAS, a reference with both transcriptome and high-density genotyping data from a small set of subjects, such as the Genotype-Tissue Expression project (GTEx), is used to build prediction models of gene expression for downstream association analyses. However, the accuracy of gene expression prediction predicted by cis-genetic variants could be compromised if these variants are located in non-regulatory elements or if they disrupt binding sites of non-transcribed TFs in target tissues (34–37). Our recent approach, sTF-TWAS (38), by integrating susceptible TF-occupied cis-regulatory elements (STFCRES) from risk-associated TFs significantly improve the detection of cancer susceptibility genes compared to the conventional TWAS approaches.

Despite the progress made by TWAS in recent years (37–43), the current models for predicting gene expression were primarily based on cis-located genetic variants (<1Mb distance), which generally account for only a modest proportion of disease heritability (44). In comparison, trans-located genetic variants may have more impact on the disease phenotype due to their advantages in population selection pressure and compensatory post-transcriptional buffering (45,46). However, including trans-located variants in TWAS analysis is a challenge due to their overwhelming numbers on gene expression than cis-located variants, requiring larger sample sizes for detection (47). Recent developed approaches,BGW-TWAS and MOSTWAS have integrated the analysis of trans-expression quantitative trait loci (trans-QTL) and demonstrated superior performance compared to S-PrediXcan and other methods (48,49). However, both approaches did not consider TF-based cell-type-specific regulatory elements to connect translocated variants with downstream target genes, despite the premise that risk TFs significantly contribute to risk loci and gene expression associated with cancer risk. Furthermore, these trans-eQTL approaches could potentially introduce false positives for co-expression genes (or TFs and genes) due to their high sequence similarity and artificial mapping (50). Therefore, an integrative TF-based epigenetic data approach (i.e., TF ChIP-seq data) is needed to prioritize trans-located variants that may play a regulatory role in gene expression.

In this work, we introduced transTF-TWAS, which included TF-linked trans-located variants, together with cis-regulatory variants for prediction model building in an effort to improve susceptible gene discovery. We showed that transTF-TWAS outperforms other methods by significantly improving prediction models and identifying disease genes. In particular, we conducted transTF-TWAS to analyze both gene expression and alternative splicing with data generated from multiple normal tissues from the Genotype-Tissue Expression (GTEx) and large-scale GWAS data for breast, prostate, and lung cancers and three brain disorders to search for disease susceptibility genes and loci (Supplementary Data 1).

### Effect of gene silencing on cell proliferation using data from CRISPR-Cas9 essentiality screens in cancer relevant cells

CRISPR-Cas9 has enabled genome-scale identification of genes that are important for the proliferation and survival of cancer cells, which have been widely used for genetic studies (28,51,52). The CERES score, a widely used metric derived from CRISPR-Cas9 essentiality screens, provides an estimation of gene dependency levels (51). We obtained the CERES score for each gene in a given cell from the DepMap portal. Subsequently, we calculated the median of negative CERES score across 45 breast-relevant cells, 8 prostate-relevant cells, and 130 lung-relevant cells. A CERES value below -0.5 serves as the cutoff to denote the essentiality of a gene for cell proliferation in CRISPR-Cas9 gene silencing experiments (29,51).

The probability mass function of the hypergeometric distribution is: 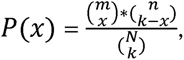 where m is the total number of genes in all cancer-related gene databases, which includes all predisposition genes, cancer drivers and CGC genes; n is the number of genes that are not included in the cancer-related gene databases (n = N – m, N= 19, 291 protein-coding genes based on the annotation from the Gencode.v26.GRCh38).

## Results

### Overview of transTF-TWAS framework

We introduced our new approach, transTF-TWAS, to build gene expression prediction models by adding TF-linked trans-located variants together with cis-variants located in STFCRES under our previous sTF-TWAS framework. Specifically, we outlined several main steps below for identifying and prioritizing trans-located variants to enhance gene expression predictions. We illustrated this methodology with the TF FOXA1 in breast cancer as an example.

Step I - Identifying TF cis-regulatory variants: we firstly identified putative TF cis-regulatory variants that potentially affect expression of a TF (e.g., FOXA1) by conducting cis-eQTL analysis and analyzing epigenetic data generated in breast-related cells. A set of the cis-regulatory variants regulating TF expression (namely TF-cis-regulatory-variants) was determined based on the significant associations between TF gene expression and genetic variants, and genetic variants, along with evidence of regulatory interactions with proximal promoters or distal enhancer-promoter regions. (Fig. 1A; Methods).

**Figure 1.**
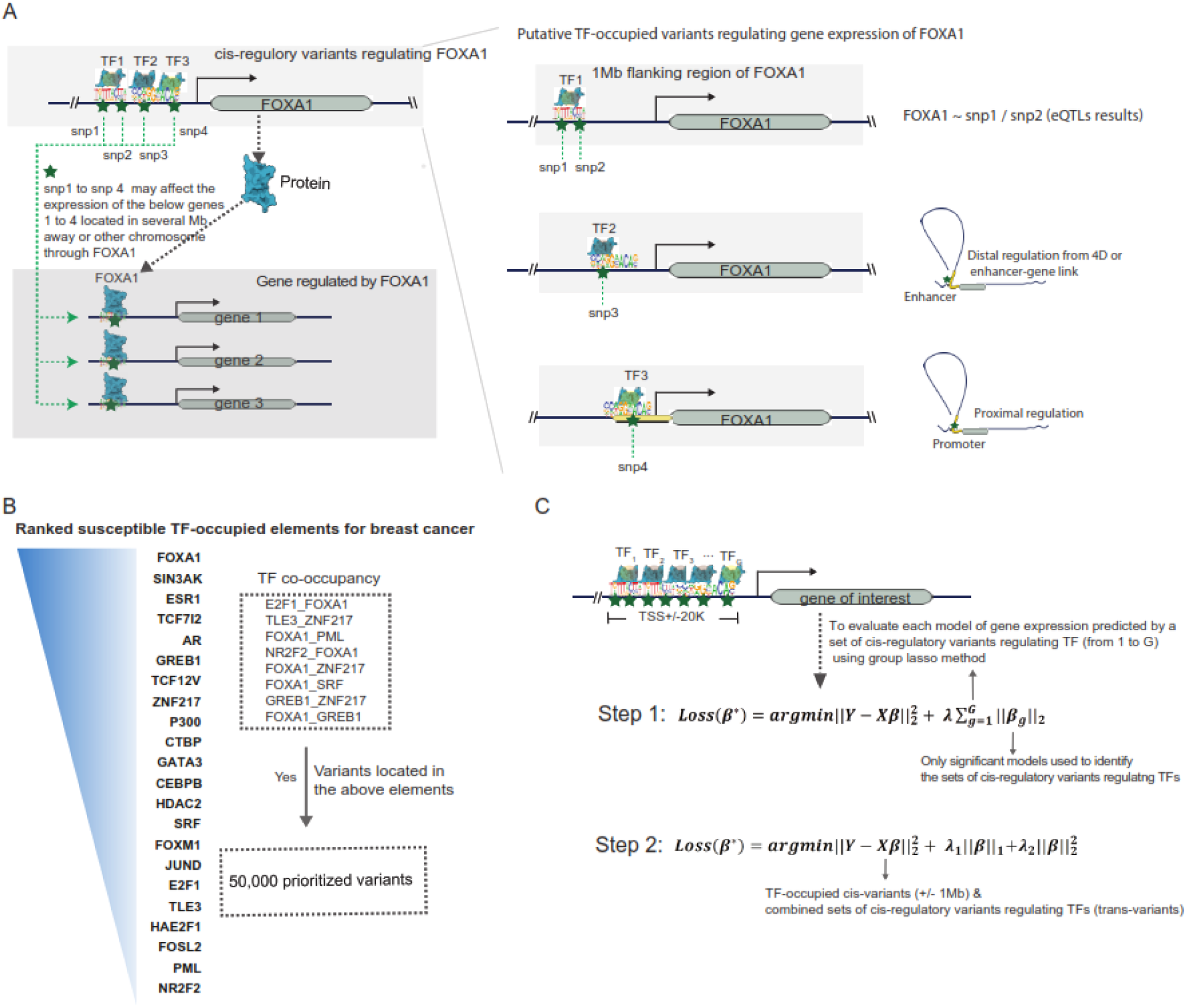
Overview of the Developed Analytical Framework. A. An illustration of how to prioritize TF-linked trans-located variants for prediction model building (using FOXA1 as an example). TF-linked trans-located variants were identified based on the as cis-eQTL (flanking 1Mb region of FOXA1, i.e., snp1 and snp2), or these variants supported with regulatory evidence through interactions with proximal promoters (i.e., snp4) or distal enhancer-promoter regions (i.e., snp3). B. Flow chart showing prioritized TF-occupied regulatory variants (50K), which were ranked based on established TF-occupied elements associated with breast cancer risk. C. An illustration of the two-step gene expression prediction model building in transTF-TWAS.

Step II - TF-gene pair discovery: we analyzed TF ChIP-seq data generated in breast cancer-related cells to characterize their genome-wide binding sites for susceptible TFs which have been identified in breast cancer from our prior work (28) (Fig. 1B; Methods). We next characterized each gene potentially regulated by all possible susceptible TFs based on the evidence of the TF-DNA binding sites that are located in its flanking of transcription start sites (TSS, +/-20K; Fig. 1C; Methods). As TF-cis-regulatory-variants have the potential to modulate the expression of the TF protein, which may result in changes in gene expression of downstream targets. Thus, these genetic variations may affect genes regulated by the TF, even if they are located several megabases away or on different chromosomes.

Step III - Model training and disease-trait association analysis: For each TF, we assessed the performance of a prediction model that utilized its TF-cis-regulatory-variants to predict expression of each target gene using Group Lasso method (i.e., number of G TFs; Fig. 1C; Methods). The Group Lasso’s property of encouraging between-group sparsity and within-group retainment aligns to our intention of selecting the actual functioning TFs and then retaining their cis-regulatory-variants. The groups survive the regularization are corresponding to those of TF-cis-regulatory-variants that may affect the expression of the gene. The final set of TF-cis-regulatory variants was identified for downstream gene expression model building by combining the groups from the significant models using standard Elastic Net (Fig. 1C; Supplementary Data1; Methods). Through expending our previous sTF-TWAS framework to build gene expression prediction models using the prioritized 50K cis-located variants (Fig. 1B), we next included TF-cis-regulatory-variants (as trans-located variants) identified from the above analysis. Here, we only focused on the set with 50K cis-located variants, as the identified genes were highly overlapped among analyses with different number of variants (i.e., 50K vs 500K variants) in our prior work (28,38). We conducted TWAS analyses by applying the gene expression prediction models, respectively, to GWAS summary statistics for breast, prostate, and lung cancers and other diseases to search for their susceptibility genes and loci (Supplementary Data 1).

### Simulation study

We conducted simulations to assess the enhancement of gene expression prediction models in statistical power by utilizing selected TF-linked translocated variants and cis-variants located in STFCRES. We compared to three existing TWAS methods (BGW-TWAS, sTF-TWAS, and S-PrediXcan), using either all cis-variants or randomly selected variants of equivalent numbers (see Methods). Initially, we validated that the type-I error of transTF-TWAS remained under control (Supplementary Figure S1). Subsequently, we examined two scenarios: causality, where genotypes influence a phenotype through gene expression intermediaries, and pleiotropy, where genotypes influence a phenotype and expression independently (Methods). In both scenarios, we observed that transTF-TWAS outperformed other TWAS methods when gene expression heritability was significantly contributed by trans-located variants, while it exhibited comparable performance to sTF-TWAS when gene expression heritability was weakly influenced by trans-located variants (Fig. 2, Supplementary Figure S2). Overall, the statistical power of transTF-TWAS increased proportionally with the heritability of both gene expression and phenotype traits and decreased with an increased number of causal variants (Fig. 2, Supplementary Figure S2). These observations offer compelling evidence that our transTF- TWAS approach, integrating TF-linked translocated variants, holds superior statistical power compared to these earlier methods.

**Figure 2.**
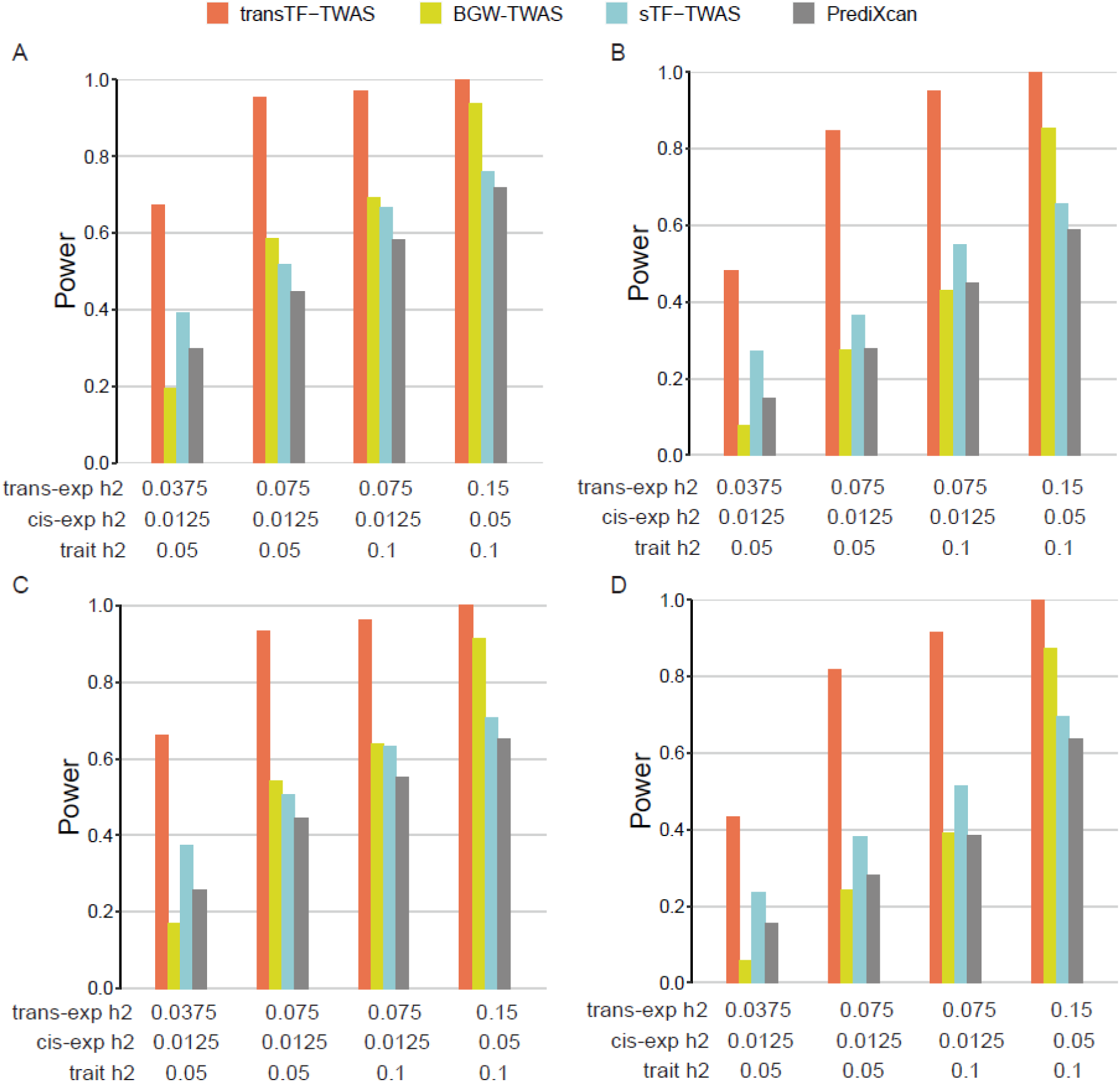
Power comparison under pleiotropy and causality scenarios for gene expression heritability that are substantially contributed by TF-linked trans-located variants. Power is indicated on the y-axis. All panels are results under an additive genetic architecture, with differing trans-variants expression heritability, cis-variants expression heritability and trait heritability denoted below each panel. A-B. under pleiotropy scenario. A. 20 causal genetic variants. B. 50 causal genetic variants. C-D. under causality scenario. C. 20 causal genetic variants. D. 50 causal genetic variants.

### Independent verification of gene expression prediction

To assess the performance of transTF-TWAS, we conducted simulations within an extension of our sTF-TWAS framework(38), termed sTF-TWAS(R), by incorporating randomly selected translocated variants of equal quantity (Methods). Initially, we compared the predictive performance of gene expression models derived from transTF-TWAS with those from sTF-TWAS(R) and sTF-TWAS. In our analysis utilizing breast tissue data from GTEx, we observed that transTF-TWAS predicted slightly more genes than sTF-TWAS(R) with an R^2^ threshold > 0.01, while it predicted over 1,800 genes more than sTF-TWAS (Supplementary Figure S3A).

Moreover, employing independent datasets generated from breast normal tissues from Kome (n=181), we demonstrated that a higher proportion of genes predicted by transTF-TWAS were validated compared to the two alternative methods (Supplementary Figure S3B). Furthermore, upon applying the prediction models to GWAS data in breast cancer, we identified 141 putative susceptibility genes using transTF-TWAS at a Bonferroni-corrected significance level of *P* < 0.05, exceeding the number identified by sTF-TWAS (62 genes) and sTF-TWAS(R) (41 genes) (Supplementary Figure S3C).

### TransTF-TWAS outperforms existing TWAS approaches in real data analysis

We next expanded our comparisons for transTF-TWAS with existing approaches including sTF-TWAS(38), PUMICE(43) and S-PrediXcan(30), in the analysis for breast, prostate and lung cancers. We showed that transTF-TWAS identified more genes than sTF-TWAS under multiple *P*-value cutoffs (Fig. 3A). As described above, we identified 141 putative susceptibility genes from transTF-TWAS, at a Bonferroni-corrected *P* < 0.05, while fewer genes were identified by sTF-TWAS (n=62), S-PrediXcan (n=52), and PUMICE (n=42) (Supplementary Figure S4; Supplementary Figure S5; Supplementary Data 2). We conducted similar comparisons for prostate and lung cancers and demonstrated consistent trends of more genes identified by transTF-TWAS compared to the other three approaches (Supplementary Figure S4; Supplementary Figure S5; Supplementary Data 3; Supplementary Data 4).

**Figure 3.**
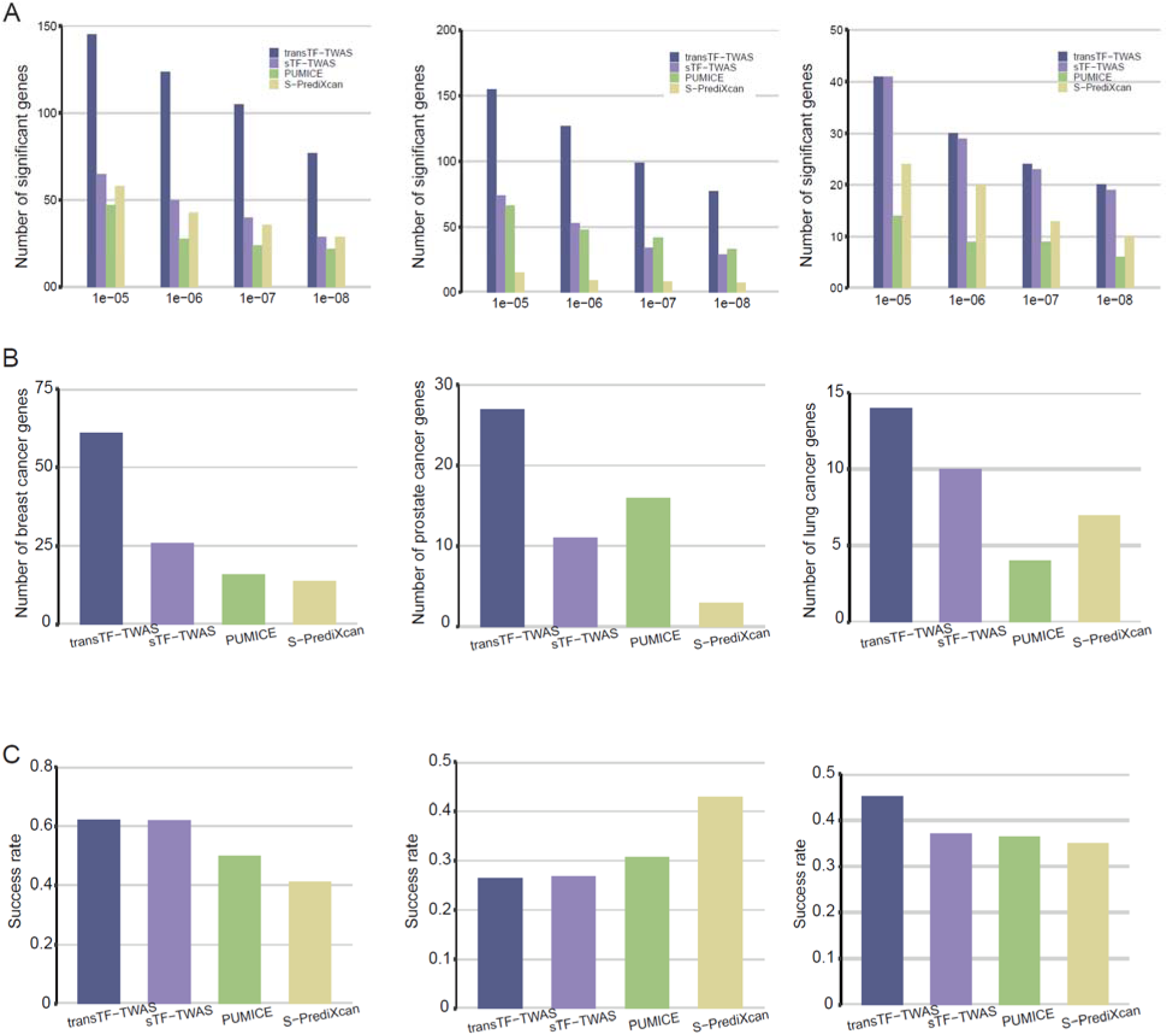
Comparison of gene-trait associations between transTF-TWAS with other TWAS approaches (sTF-TWAS, PUMICE, and S-PrediXcan) for breast, prostate and lung cancer. A. Bar chart showing the number of genes identified from transTF-TWAS and other TWAS approaches under various *P*-value cutoffs (i.e., *P* < 1e-05, 1e-06, 1e-07, and 1e-08). The P-values are the nominal P-values from the Z score test from TWAS. B. Bar chart showing a comparison between the total number of target cancer related genes among transTF-TWAS and other TWAS approaches. C. Bar chart showing a comparison of the proportion (success rate) of target cancer related gene among transTF-TWAS and other TWAS approaches, relative to the total number of genes identified from the set.

Next, we utilized knowledge-based disease-relevant genes to evaluate the potential false positives of our gene-trait associations identified from transTF-TWAS compared to other approaches. We conducted functional annotation for the genes identified by transTF-TWAS and sTF-TWAS with known target cancer-related genes of interest (Methods). Our analysis revealed that transTF-TWAS detected more breast cancer-related genes (n=61) compared to sTF-TWAS (n=26) (Fig. 3B), with a comparable proportion (62.2% for transTF-TWAS vs. 61.9% for sTF-TWAS) (Fig. 3C). Additionally, our approach identified an overall higher quantity and a higher or comparable proportion of known cancer-related genes for both prostate and lung cancers compared to other approaches (Fig. 3B,C).

### Genetically driven key regulators and their associated networks underlying cancer risk

We showed that transTF-TWAS detected more genes than sTF-TWAS in breast, prostate, and lung cancers, whereas a large number of significant genes were uniquely detected by transTF-TWAS (Fig. 4A). To further illustrate how these unique genes contributed by trans- located variants, we examined whether these genes can be predicDataby sTF-TWAS. We found that most of the unique genes in breast and prostate cancers failed to be genetically predicted by sTF-TWAS, indicating the trans-located variants significantly contributed risk gene discovery via the improved gene expression prediction performance (Fig. 4A; Supplementary Data 5; Supplementary Data 6; Supplementary Data 7).

**Figure 4.**
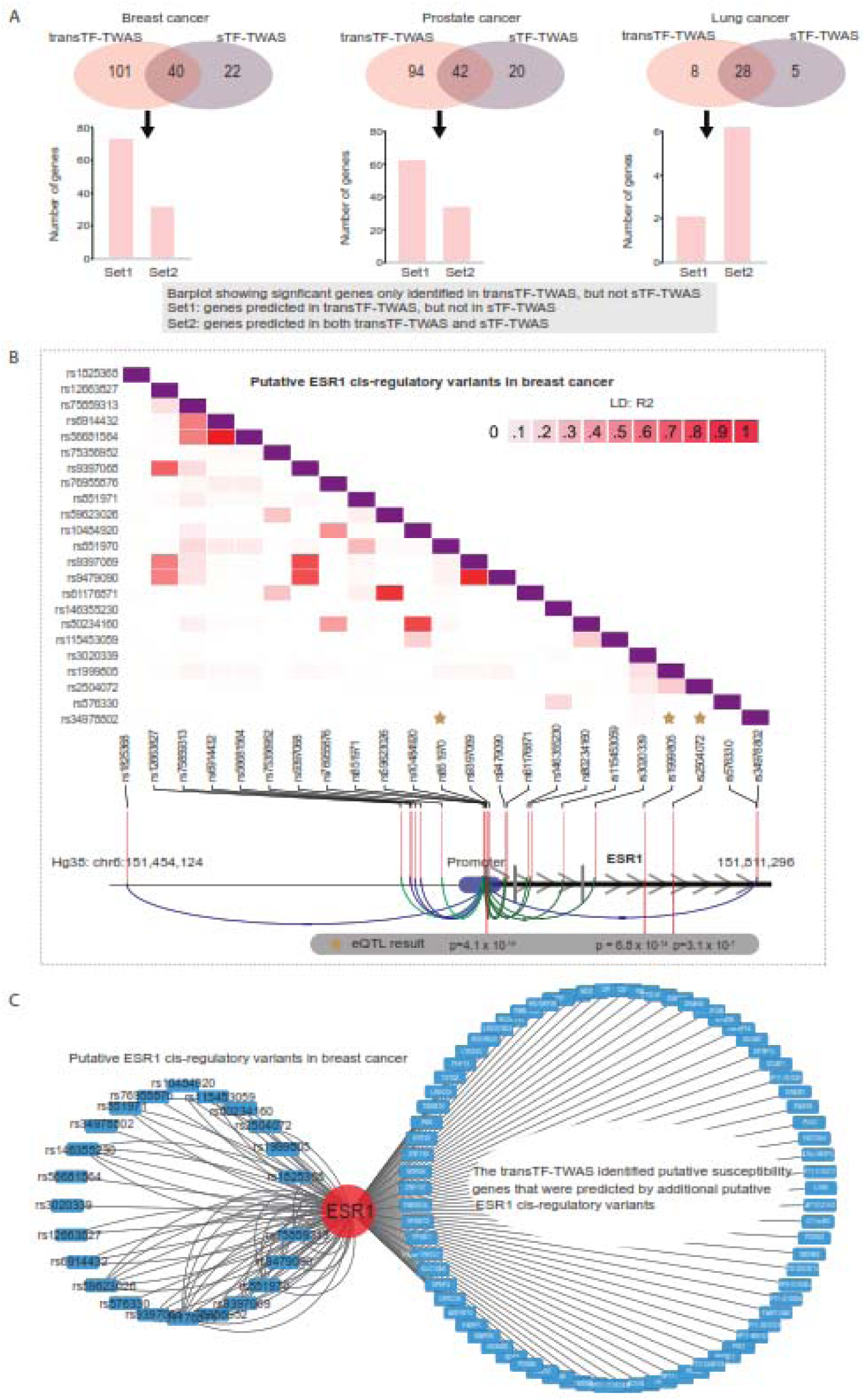
The gene regulatory network underlying cancer risk driven by master regulators. A. Venn diagrams showing the number of putative susceptibility genes commonly or uniquely identified by transTF-TWAS and sTF-TWAS. An arrow points from the uniquely identified genes by transTF-TWAS to a bar chart showing: Set 1: genes predicted in transTF-TWAS, but not in sTF-TWAS. Set 2: genes predicted in both transTF-TWAS and sTF-TWAS. B. A heatmap showing the LD structure among putative ESR1 cis-regulatory variants in breast cancer. The ESR1 cis-regulatory variants are trans-located variants that present the strongest associations with cancer risk in the prediction model. C. A network showing the connections between the putative ESR1 cis-regulatory variants in breast cancer and putative susceptibility genes identified by transTF-TWAS that were contributed by putative ESR1 cis-regulatory variants.

Of the identified genes, we next evaluated the lead trans-located variants that present the strongest associations with cancer risk in the prediction model for each of our identified putative susceptibility genes. In breast cancer, we observed that the lead variants are significantly enriched in the TF-cis-regulatory-variants for ESR1 (n=73 genes), followed by TCF7L2 (n=13), and FOXA1 (n=11) (Fisher’s exact test, *P* < 0.01 for all; Fig. 4B, C; Supplementary Data 8; Supplementary Data 9). In prostate cancer, we observed that the lead variants are significantly enriched in NKX3-1 (n=61 genes), followed by GATA2 (n=13) (Fisher’s exact test, *P* < 0.01 for all; Supplementary Data8; Supplementary Data 10). These results highlighted these genetically-driven key regulators and their associated regulatory networks that underlie cancer susceptibility.

### Charactering novel cancer risk genes identified by transTF-TWAS

To further characterize putative susceptibility genes and loci identified under our transTF-TWAS framework (Fig. 5), we additionally analyzed alternative splicing (sp-transTF- TWAS; see Methods) for breast, prostate and lung cancers (Supplementary Figure S6; Supplementary Data 11; Supplementary Data 12; Supplementary Data 13). We comprehensively compared our findings from both transTF-TWAS and sp-transTF-TWAS, with those reported from previous TWAS, eQTL, or other genetic studies for breast (32,53)^;^ (19)^;^ (28,38,54,55), prostate (31,38,56–58) and lung cancers (38,58,59).

**Figure 5.**
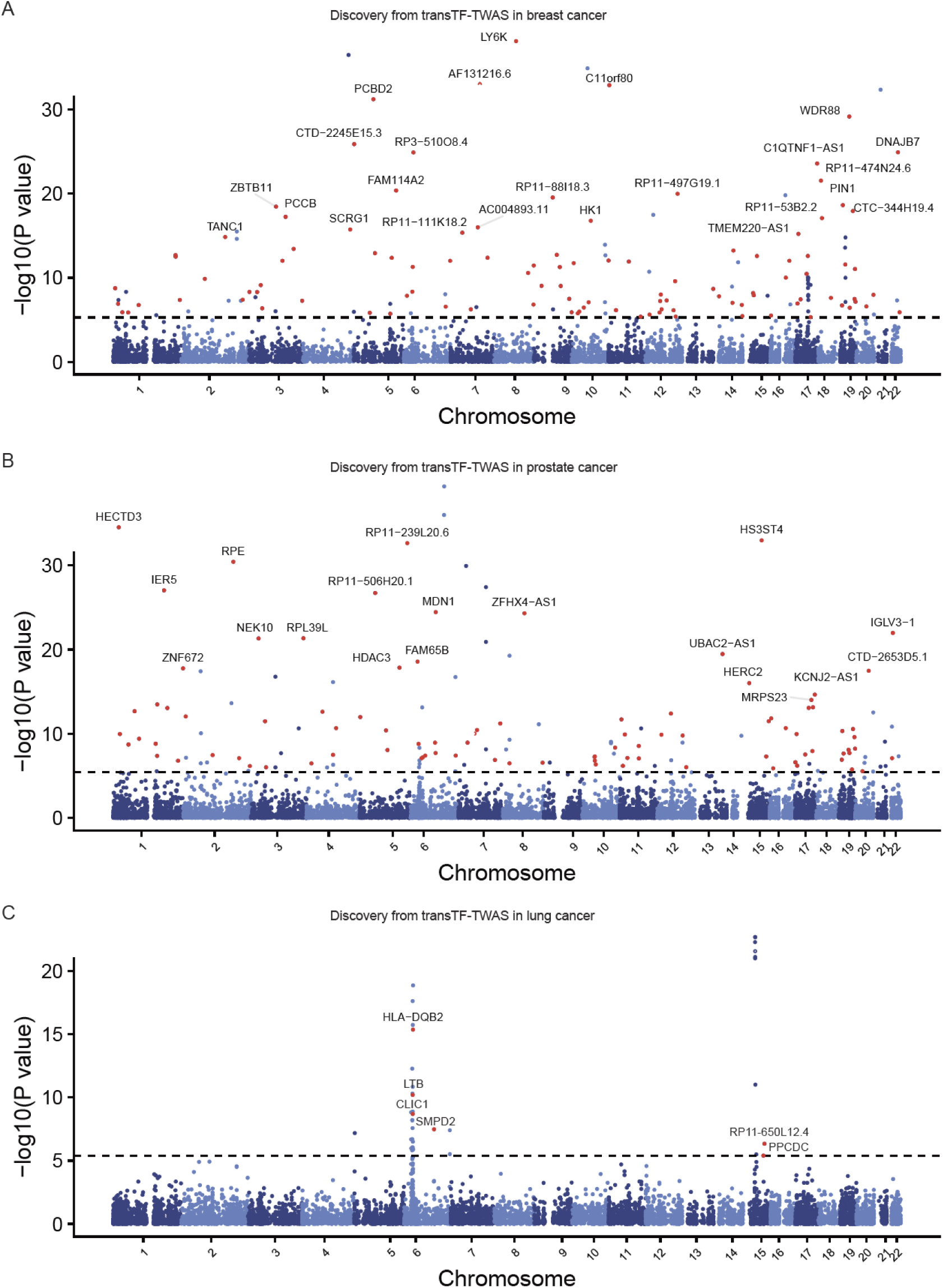
Putative susceptibility genes identified by transTF-TWAS. Manhattan plots showing the associations identified from transTF-TWAS. Red dots indicated all newly identified susceptibility genes, and the grey dashed line refers to Bonferroni-corrected *P* < 0.05. The newly identified putative susceptibility genes with P < 10^-15^ were highlighted. The *P*-values are the raw *P*-values from the Z score test from TWAS (two-sided). A. breast cancer. B. prostate cancer. C. lung cancer.

For breast cancer, we identified 141 putative susceptibility genes from transTF-TWAS and 239 putative susceptibility genes from sp-transTF-TWAS, at a Bonferroni-corrected *P* < 0.05. Combing the results from both analyses, we identified 374 putative breast cancer susceptibility genes, including 212 genes at 163 novel loci (more than 1Mb away from any previous GWAS-identified risk variant for breast cancer) and 53 previously unreported located in GWAS loci (Fig. 6A, B; Supplementary Data14). For prostate cancer, we identified 136 putative susceptibility genes from transTF-TWAS and 318 putative susceptibility genes from sp-transTF-TWAS. Combing the results from both analyses, we identified 443 putative prostate cancer susceptibility genes, including 251 genes at 193 novel loci and 75 genes previously unreported located in GWAS loci (Fig. 6A, B; Supplementary Data 15). For lung cancer, we identified 36 putative susceptibility genes from transTF-TWAS and 41 putative susceptibility genes from sp-transTF- TWAS. Combing the results from both analyses, we identified 70 putative lung cancer susceptibility genes, including 2 genes at one novel locus and 9 genes previously unreported located in GWAS loci (Fig. 6A,B; Supplementary Data 16). Taken together, our analysis revealed a total of 887 putative susceptibility genes for these three cancer types, including 137 that were previously unreported in GWAS loci and 465 in loci unreported by GWAS (Supplementary Data 17; Supplementary Data 18).

**Figure 6.**
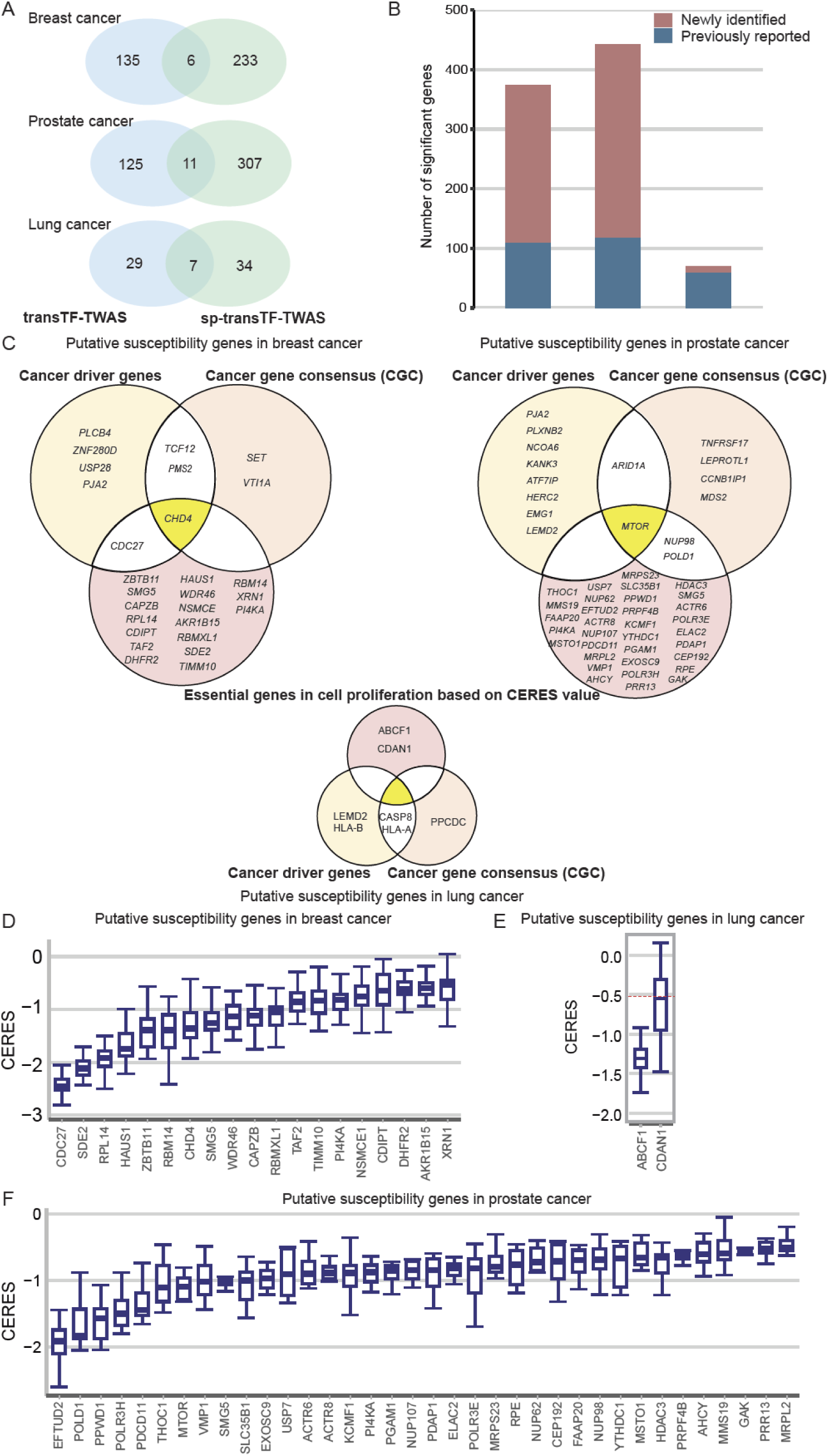
Putative susceptibility genes identified by transTF-TWAS and sp-transTF-TWAS. A. Venn diagrams showing the number of putative susceptibility genes commonly or uniquely identified by transTF-TWAS and sp-transTF-TWAS. B. Bar chart showing the total identified putative susceptibility genes combined from transTF-TWAS and sp-transTF-TWAS for breast, prostate and lung cancer. C. Venn diagrams showing all newly identified genes that were cancer driven genes, Cancer Gene Census (CGC), or genes with CERES<-0.5 for breast cancer, prostate and lung cancer. D-F. Boxplot showing all newly identified genes with evidence of essential roles in cell proliferation based on a cutoff of median CERES values < -0.5 for D) breast cancer (sample size: 45 cell lines), E) lung cancer (sample size: 130 cell lines), and F) prostate cancer (sample size: 8 cell lines). In the boxplots shown in these figures, the whiskers denote the range; the boxes denote the interquartile range; the middle bars in denote the median.

### Functional evidence of oncogenic roles for the identified putative susceptibility genes

We next examined whether our identified putative cancer susceptibility genes had been reported as predisposition genes(60,61), cancer drivers (62,63), or Cancer Gene Census (CGC) genes (64) (Methods). We found eight cancer driver genes and five CGC among previously unreported genes for breast cancer, as well as six cancer driver genes and eight CGC among previously reported genes (Fig. 6C). Similarly, for prostate cancer, we found ten cancer driver genes and eight CGC among previously unreported genes, and six cancer driver genes and four CGC among previously reported genes (Fig. 6C). For lung cancer, we identified four cancer driver genes and three CGC among previously reported genes. Functional enrichment analysis showed that our identified genes were significantly enriched in those known cancer-related genes with *P* = 0.0044 for breast cancer, *P* = 0.0097 for prostate cancer and *P* = 0.012 for lung cancer (Methods).

We also explored the functional roles of the identified putative susceptibility genes using CRISPR-Cas9 screen silencing data to investigate gene essentiality on cell proliferation in breast (n=45), prostate (n=8), and lung (n=130) cancer relevant cell lines (Methods). Using a cutoff of median CERES Score < −0.5 in the above cells, following the previous literature (51,52), we provided strong evidence of essential roles in cell proliferation for 19 previously unreported genes for breast cancer (Fig. 6D); 36 unreported genes for prostate cancer (Fig. 6E); and two unreported genes for lung cancer (Fig. 6F).

### TransTF-TWAS strengthens non-cancer risk gene discovery

To evaluate the generalizability of transTF-TWAS, we conducted additional analysis for brain disorders including schizophrenia (SCZ), Alzheimer’s disease (AD), and autism spectrum disorder (ASD). By comparison, we also conducted S-PrediXcan and sTF-TWAS for each of the diseases. We found that transTF-TWAS identified more putative susceptibility genes than both sTF-TWAS and S-PrediXcan for AD and ASD. Using ASD as an example, we identified eight putative susceptibility genes from transTF-TWAS at a Bonferroni-corrected *P* < 0.05, while only one and six genes were identified by S-PrediXcan and sTF-TWAS, respectively. The results suggest that our transTF-TWAS approach has broad applicability for enhancing the discovery of disease susceptibility genes (Supplementary Figure S7).

## Discussion

In this study, we demonstrated that the new approach, transTF-TWAS, significantly improved the detection of putative cancer susceptibility genes with increased statistical power and accuracy over other existing TWAS approaches (i.e., sTF-TWAS (38), BGW-TWAS (48), PUMICE (43), and S-PrediXcan (30)). Under transTF-TWAS framework, we predicted alternative splicing and gene expression and applied these models to large GWAS datasets for breast, prostate, and lung cancers. Our analysis revealed total 887 putative cancer susceptibility genes, including 465 in regions not yet reported by previous GWAS and 137 in known GWAS loci but not yet reported previously. Many of the newly identified associations have been supported by their oncogenic roles in cancer development (62–64), including 88 cancer driver genes, CGC or those with strong evidence of essential roles in target cancer cell proliferation. These findings provide new insights into the genetic susceptibility of the three common cancers.

Previous TWAS mainly use cis-genetic variants in building gene expression models. However, investigation into trans-located variants has been limited due to statistical analysis burden of their large numbers. To address this, transTF-TWAS is to identify TF-linked trans- located variants by comprehensively characterizing TF-cis-regulatory-variants and using Group Lasso to select a set of these variants to significantly contribute prediction models. The use of Group Lasso can be powerful in identifying a set of TF-cis-regulatory-variants, which may affect the expression of the TF regulated target genes. Our analysis identified many TF-linked trans- located variants (i.e., average 10 for each gene for breast cancer) contributed to gene expression prediction, which included several thousand newly predicted genes missed by other approaches. In prior research, it has estimated that 60%∼90% of heritability of gene expression can be explained by distal genetic variation in various tissues (65). We additionally analyzed expression predictions for a total of 4,625 genes that can be predicted by both TF-linked translocated and cis-located putative regulatory variants using breast normal tissues from the GTEx. We found that translocated variants exhibit a greater heritability contribution to gene expression compared to cis-variants (Supplementary Figure S8). We also conducted additional analysis of trans-variants (included in our gene expression prediction models) from trans-eQTL analysis in target tissues (i.e., breast normal tissues) and non-target tissues (i.e., non-breast normal tissues) using data in GTEx. We showed that the absolute beta values are significant higher in target tissues than non-target tissues (0.228 vs 0.099 vs 0.056; *P* < 0.05; Supplementary Figure S9). These results provide additional support that the model building for a large majority of genes by introducing TF-linked trans-located variants can facilitate putative causal susceptibility gene discovery via such TF-medicated mechanisms.

Much efforts (20,21,27), including our work (28), have established cancer susceptible TFs, whose DNA binding sites altered by risk genetic variants that regulate cancer susceptibility genes. However, it remains unclearly how susceptibility TF-based transcriptional networks underlying genetic susceptibility to common cancers. In this study, transTF-TWAS can strengthen susceptibility gene discovery through integrating the prior information of TF-cis-regulatory-variants altered regulators and their downstream target genes. For breast cancer, we observed that the variants potentially regulated TFs significantly contribute to expression prediction of their downstream regulated genes. In turn, the putative susceptible genes that were identified appear to be commonly regulated by FOXA1 and ESR1 through the upstream genetically-driven regulatory mechanisms, further highlighting their key roles in driving breast cancer susceptibility (Fig. 3; Supplementary Data9). Similarly, we also highlighted key TFs, NKX3-1 (66–69) and GATA2 (70–73) for prostate cancer (Supplementary Data10).

Unfortunately, we did not observe significant TFs in lung cancer, likely due to the less genetic effects of the TF on downstream regulated genes (38).

Our findings are in line with the evidence that trans-located genetic variants may have more impact on diseases than cis-located genetic variants(48,65,74). For our transTF-TWAS, one critical step is to prioritize TF-linked trans-located variants for model building based on identifying TF-cis-regulatory-variants and TF regulated downstream genes. The concept of our approach can also be used to identify trans-located variants based on long distance-based epigenetic signals (>1Mb) such as distal chromatin-chromatin interaction, enhancer-gene link, enhancer-gene correlation as well as trans-eQTLs (75–78). Thus, our transTF-TWAS will further strengthen disease susceptibility gene discovery with increasing availability of extensive epigenetic datasets in future studies. On the other hand, the prior information of TF-linked trans- located variants can be integrated into other extensions of TWAS, such as multiple-tissue approaches (UTMOST (79) and S-MultiXcan (80)), or variance component (Kernel) TWAS (39,40) or instrumental-variable approaches (41,42).

In conclusion, we demonstrated that our transTF-TWAS, by integrating TF-linked trans- located variants with TWAS, significantly improved disease susceptibility gene discovery and advanced our understanding of complex human diseases, including cancers. Our study also highlighted several genetically driven key regulators and their associated regulatory networks underlying disease susceptibility.

## Methods

### Data resources

We obtained the individual-level genotype dataset from GTEx (v8)(81,82), which was quality controlled using PLINK(83). Summary statistics of GWAS data for breast cancer were obtained from the Breast Cancer Association Consortium (BCAC), which has generated GWAS data for 122,977 cases and 105,974 controls from European descendants. GWAS data for prostate cancer were released from the European descendants from the Prostate Cancer Association Group to Investigate Cancer Associated Alterations in the Genome (PRACTICAL)(84), with 79,194 cases and 61,112 controls from European descendants. GWAS data for lung cancer were obtained from the websites of the Transdisciplinary Research of Cancer in Lung of the International Lung Cancer Consortium (TRICL-ILCCO) and the Lung Cancer Cohort Consortium (LC3) (85), with 29,266 cases and 56,450 controls from European descendants. GWAS summary statistics for schizophrenia (SCZ, N= 70,100), Alzheimer’s disease (AD, N=22,246), and autism spectrum disorder (ASD, N= 10,263) were downloaded from the Psychiatric Genomics Consortium website (PGC).

The TF-occupied regulatory variants for breast, prostate, lung cancers, and three brain disorders were collected based on ChIP-seq data of transcription factors (TFs) generated in diseases related cell lines from the Cistrome database(86). We evaluated their quality control based on the guidance from the database and selected high-quality datasets for downstream analysis. Detailed ChIP-seq data for breast, prostate, lung cancers and brain disorders were described in our previous work (28,38).

We included germline whole genome sequencing (WGS) and RNA-sequencing (RNA-seq) data from GTEx (release 8) (81,82) for normal breast tissue, prostate tissue, lung tissue, and brain cortex tissue. We selected tissue samples from 151 women for breast tissue, 221 men for prostate tissue, 515 individuals for lung tissue, and 205 individuals (both sexes) for brain cortex tissue. The fully processed, filtered, and normalized gene expression data matrices (in BED format) were downloaded from the GTEx portal. The WGS file and sample attributes were obtained from dbGaP, and the subject phenotypes for sex and age information were obtained from the GTEx portal. The covariates used in eQTL analysis were obtained from GTEx_Analysis_v8_eQTL_covariates.tar.gz, and the covariates for sQTL analysis were obtained from GTEx_Analysis_v8_sQTL_covariates.tar.gz, both of which were downloaded from the GTEx portal. Normal breast tissue samples for both RNA-sequencing and genotyping from 181 individuals of European were collected through the Susan G. Komen Normal Tissue Bank (Kome). Genotype and gene expression data generation and processing have been described in our previous sTF-TWAS work (38).

We downloaded approximately 3.6 million DNase I hypersensitive sites (DHSs) regions within human genome sequence (87). The enhancers regions were downloaded from EpiMap repository (88), which contains ∼2M non-tissue specific enhancers regions. The CAGE peak regions were downloaded from FANTOM5 (89), and we also included all regions within transcription start site (TSS) +/-2K for each gene as promoter regions. The eQTLs were downloaded from the GTEx portal (81,82) and eQTLGen (90). The Enhancer to gene link information across 833 cell-types were downloaded from EpiMap repository (88). We all used cell-type specific chromatin-chromatin interaction data from the 4D genomics and previous literature (91,92).

To analyze cancer-related susceptibility genes, we downloaded a list of gene sets from the Molecular Signatures Database (MGB) on Gene Set Enrichment Analysis (GSEA).

Additionally, we downloaded lists of predisposition genes from previous literatures (61,93), cancer-driven genes from two previous literatures (94,95), and CGC (96) from the COSMIC website. To investigate the effect of an individual gene on essentiality for the proliferation and survival of cancer cells, we downloaded two comprehensive datasets, "sample_info.csv" and "CRISPR_gene_effect.csv," from DepMap Public 21Q4.

### Identifying TF-cis-regulatory-variants

To determine a set of the cis-regulatory variants that potentially regulate TF expression (namely TF-cis-regulatory-variants), we first prioritized putative regulatory variants by only including TF-occupied variants that are located in DNase I hypersensitive sites (DHSs) (87), enhancer regions(88) and promoter regions (88). Of them, the significant associations between a TF and its cis-genetic variants were identified at a nominal p-value < 0.05, based on the eQTL analysis in both target tissues and whole blood samples using data from GTEx portal(81,82) and eQTLGen(90). Furthermore, we also analyzed epigenetic data to search regulating evidence by these variants through interactions with proximal promoters or distal enhancer-promoter regions. Specifically, we examined if these variants are located in the promoter region of a TF (TSS +/- 2K) or enhance region with an evidence of the enhancer linking to the TF based on expression- enhancer activity correlation across 833 cell-types from the EpiMap repository (88), as well as chromatin-chromatin interaction data from the 4D genomics and previous literature (91,92).

Finally, the TF-cis-regulatory-variants were identified based on the significant associations from eQTL results, and the regulatory evidence from the variants linked to the TF.

### Gene expression prediction model building based on trans-located variants

We analyzed TF ChIP-seq data generated in target cancer-related cells to characterize their genome-wide binding sites for susceptible TFs using data from the Cistrome database (86). We next characterized each gene potentially regulated by all possible susceptible TFs based on the evidence of their TF-DNA binding sites that are located in its flanking 20Kb of TSS (i.e., number of *G* TFs). For each TF, we assessed the performance of a prediction model that utilized its TF-cis-regulatory-variants to predict expression of each target gene using Group Lasso method. We trained a Group Lasso to select a group of TF-cis-regulatory-variants from each TF (i.e., 1 to G TF).

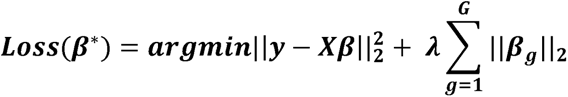

where the coefficient in ***β*** are divided into *G* groups and ***β****_g_* denotes the coefficients vector of variants in the *g*-th group. ***X*** are all trans-located variants from *G* groups. y is normalized gene expression data generated in different tissue samples from GTEx v8. In Group Lasso, the regularizor, ||***β****_g_*||_2_, also called *l*_2,1_-norm consists of the intra-group non-sparsity via *l*_2_-norm and inter-group sparsity via *l*_1_-norm. Only significant models were used to determine those groups of TF-cis-regulatory-variants that may affect the expression of the gene. The final set of TF-cis-regulatory variants was identified for downstream gene expression model building by combining the groups from the significant models. We next built gene expression prediction models for the final sets of TF-cis-regulatory variants and cis TF-occupied variants using standard Elastic Net under our sTF-TWAS framework. For each gene, the gene expression level was regressed on the number of effect alleles (0, 1, or 2) for each genetic variant with adjustment for top 5 genotyping PCs, age, and other potential confounding factors (PEERs). We used 30 PEER factors for our downstream model building based on the recommendation for breast, prostate and brain tissues, and 60 PEER factors for lung tissue. Prediction model performance was assessed using the R^2^ via a 10-fold cross-validation.

### Simulation study and external verification to assess gene expression predictions

To evaluate prediction performance of our developed approach, we simulated scenario for each gene that had the equal number of artificial TF groups with our transTF-TWAS. We also randomly generated the same number of trans-located genetic variants (> 1Mb distance) within each TF group with our transTF-TWAS. Similarly, we next used Group Lasso to select significant groups from the artificial TF groups. The final set of trans-located variants was identified for downstream gene expression model building by combining the groups from the significant models. We next built gene expression prediction models for the final sets of and cis TF-occupied variants under sTF-TWAS framework. The models of genetically predicted gene expression were built in breast normal tissues from the GTEx project. To externally verify gene expression prediction performance, we first used the same analytical protocol to build the prediction models using standard Elastic Net based on normalized gene expression data generated in breast tissue from the GTEx (v8), and then we re-calculated the prediction performances in terms of variance explained (R^2^) using selected variants trained from the GTEx based on an independent dataset generated in breast normal tissues from Kome, where the genotype and gene expression data were processed following the protocol in GTEx.

### Association analyses between predicted gene expression and cancer risk

To evaluate associations of genetic predicted gene expression with cancer risk, we applied the weight matrix obtained from the gene prediction models to the summary statistics implemented in S-PrediXcan (97). The statistical method described in the following equation that was also described elsewhere(32,33), was used for association analyses.

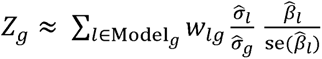

Here, Z-score was used to estimate the association between predicted gene expression and cancer risk. Here, *w_lg_* is the weight of genetic variant *l* for predicting the expression of gene

*g*·*β̂_l_* and se(*β̂_l_*) are the GWAS-reported regression coefficients, and its standard error for variant *l*, and *σ̂_l_* and *σ̂_g_* are the estimated variances of variant *l* and the predicted expression of gene *g*, respectively.

By comparison, we also performed TWAS analysis using PUMICE(43) (Prediction Using Models Informed by Chromatin conformations and Epigenomics) with default settings. PUMICE improves the accuracy of transcriptomic imputation through utilizing tissue-specific 3D genomic and epigenomic data to prioritize regions that harbor cis-regulatory variants. The source codes of PUMICE were obtained from https://github.com/ckhunsr1/PUMICE. The precomputed models trained in breast, prostate and lung tissues from GTEx v8 can be found under https://github.com/ckhunsr1/PUMICE/tree/master/models_GTEx_v8.

### Simulation study to assess type-I error

We conducted the null simulation to evaluate the type-I error of our transTF-TWAS. The individual-level genotype data provided by the GTEx was used as the reference dataset to simulate the gene expressions and the 1000 Genomes Project dataset was used to simulate phenotype and conduct GWAS/TWAS analyses. We first randomly generated phenotype values (0 or 1) independently from the genotype. We then conducted logistic regression analysis to generate the GWAS summary statistics using the phenotype values and the genotype data from the 1000 Genomes Project. We next randomly generated putative TF-occupied regulatory variants and TF-linked trans-located variants for model training. The protocol for randomly selected these putative TF-occupied regulatory variants are the same as our previous sTF-TWAS work (38). Specifically, to generate 50K cis-located variants occupied by an artificial TF, we prioritized a set of variants using generalized linear mixed models to analyze GWAS summary statistics of all genetic variants and the TF’s binding status. Specifically, we randomly assigned 50K TF-occupied variants to a value “1” and the remaining variants to a value “0” (i.e., 1 for a variant located in a TF binding site, 0 otherwise). We then used generalized linear mixed models to estimate an association between the Chi-squared values (Y) and TF binding status of genetic variants. We prioritized a set of variants based on the association for a given ‘TF’ with cancer risk at *P* < 0.05. We repeated the above statistical analysis (i.e., >4000 times) and used prioritized sets of variants as TF-occupied cis-regulatory variants together with the randomly selected trans-located variants for our downstream model training. To generate a set of TF-linked trans-located variants: We randomly selected 50 cis-located variants as eQTL variants for an artificial TF. We repeated the process to generate 10 sets of cis-located variants for 10 artificial TFs. Theses TFs are all considered as TF-gene pairs for downstream model training. The Group Lasso was used to train each set of cis-located variants for each TF in the gene expression prediction model to examine if the TF significantly contributes to the prediction (i.e., non-zero coefficients). For the groups surviving the regularization, they were considered as trans-located variants along with TF-occupied cis-regulatory variants are all included in the Elastic Net to train and prioritize genetic variants to predict gene expression. We further conducted TWAS analysis based on the well predicted gene expression models (R^2^ > 0.01) and GWAS summary statistics. We repeated the above simulation processes 100 times to increase the robustness.

### Simulation study to assess statistical power

The process of simulation is structured into three components: (1) high-level genetic architectures, i.e., causality and pleiotropy; (2) trait heritability and expression heritability; (3) expression heritability contributed by cis- and trans-variants. They are described below:

We conducted simulations under two representative high-level architectures: (1) causality where genotype causes phenotypic changes via the mediation of gene expression, and (2) pleiotropy where genotype contributes to phenotype and gene expression independently. To simplify simulations, under both scenarios, we simulated gene expressions and phenotypes using an additive genetic architecture. Under the additive architecture, phenotypes and gene expressions are simulated by the sum of genetic effects:

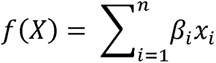

Where *X* is the genotype matrix from either the GTEx or 1000 Genomes Project, *X* = {*x*_1_, *x*_2, …_*x_n_*}. The effect size *β_i_* is drawn from the standard normal distribution N (0,1), which will be used in the downstream TWAS analysis. The formula for simulating gene expression:

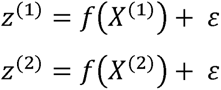

Where *z*(^1^) is the gene expression simulated using genotype from the GTEx (*X*(^1^)) and *Z*(^2^) is the gene expression simulated using genotype from the 1000 Genomes Project (*X*(^2^)). Here the super-index (1) indicates that the data is from the expression dataset, GTEx, whereas the super- index (2) indicates the data is from the GWAS dataset, which is the 1000 Genomes Project in this simulation.

The formula for simulating phenotype under the causality scenario:

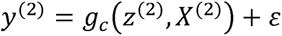

Where *y*(^2^) is the phenotype simulated with *g_c_* function, where.*g_c_*(*z*(^2^),*X*(^2^)) = Z(^2^)+ε. The formula for simulating phenotype under the pleiotropy scenario:

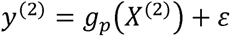

Where y(^2^) is the phenotype simulated with *g_p_* function, where *g_p_*(*X*(^2^)) employs the same format to *f*(*X*(^2^)), except that the variance component is rescaled by gene expression heritability instead of trait heritability.

The above genetic architectures define how genetic components contribute to each phenotype. Using the genetic components, we generated phenotypes where the variance component attributed to genotype, or heritability, equals a preselected value h^2^. That is, given the variance of the phenotype’s genetic component as 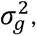 we solved 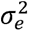 to satisfy that 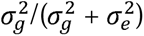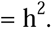 We then sampled from the normal distribution 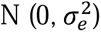 to determine the strength with which nongenetic components such as noise or environmental effects contribute to phenotype. Finally, the sum of the genetic and nongenetic components were used as the simulated phenotype in association mapping and power calculations.

To simulate the effect of TF-occupied cis-regulatory variants, we followed the same procedure as we did for sTF-TWAS. For each target gene, we first randomly selected 200 variants from the local gene regions (+/- 1Mb of the gene body) as potential predictors. When simulating gene expression heritability contributed by trans-located variants: we randomly selected 5 (or 10) genetic variants with minor allele frequency (MAF) larger than 1% from these 200 variants as the functional causal cis-located genetic variants. In addition, we simulate the effect of trans-located genetic variants. We first randomly selected 10 regions across the whole genome (> 1Mb of the gene body), and each region containing 50 randomly selected genetic variants as 10 trans-located variants group. Among these 10 regions, we randomly select 3 (or 4) regions as the real functional trans- region, and among each trans- region, we randomly select 5 (or 10) genetic variants as the real causal trans-located genetic variants. In total, we simulated 20 (or 50) real causal genetic variants for each target gene. The relative variance components contributed between cis- and (aggregated) trans- genetic variants are then re-scaled to the desired ratio using a similar re-scaling model that structures the relative contribution between genetic contribution 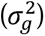 and noise 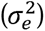 described above.

To illustrate the improvement of statistical power of transTF-TWAS which incorporates prior knowledge of the potential trans-located variants group, we compared the statistical power of transTF-TWAS with two other TWAS approaches only using cis-genetic variants: 1) PrediXcan: using all cis-genetic variants (+/-1Mb of the gene body); 2) sTF-TWAS: using 200 randomly selected genetic variants; and one TWAS approach also including trans-genetic variants for predictive model building: 3) BGW-TWAS: using all genetic variants across the whole genome.

For all models, the gene expressions are simulated using genotype data from GTEx, and phenotypes are simulated using genotype from the 1000 Genomes Project. This simulated the situations where the real gene expressions are not available in the target dataset (represented by the 1000 Genomes Project genotype and simulated phenotype); instead, we used another reference dataset (represented by the GTEx genotype and simulated expressions) to train the weights for each gene. For each of the genetic architectures and their associated parameters, we simulated **1,000 datasets,** in which causal variants were randomly selected. We then test each protocol’s ability to successfully identify the genes in each dataset, where success was defined as a Bonferroni-corrected P-value that is lower than a predetermined critical value (0.05).

### Genetically-driven key regulators and their associated networks regulating breast cancer susceptibility genes

For each of the identified putative susceptibility genes, we evaluated the lead variant that present the strongest associations with cancer risk in the prediction model. If the lead variant was a trans-located variant, we next identified its potential regulated TF based on the previous analysis of TF-cis-regulatory-variant (see the preceding section). A TF-gene pair was further determined based on the above information of the lead trans-located variant linked to both the gene and TF. Based on the information of TF-gene pairs, a TF-transcriptional network was built using Cytoscape 3.9.1(98). To determine whether our identified susceptibility genes show significant enrichment in a TF (acting as an up-regulator based on TF-gene pairs) of interest, we compared this TF to the remaining combined TFs as background using Fisher’s exact test. This statistical test employs a 2x2 Data to ascertain the significance of enrichment for the TF of interest.

### Sp-transTF-TWAS analysis

In our transTF-TWAS, we incorporated both putative cis-regulatory variants located in STFCRES and TF-linked translocated variants for gene expression prediction and downstream association analysis. As an extension of transTF-TWAS with a broader genetically regulatory mechanism (e.g., eQTL vs. alternative splicing QTL), we also utilized both putative cis-regulatory variants in STFCRES and TF-linked translocated variants for predicting alternative splicing expression. For those genetically predicted well alternative splicing (R^2^ > 0.01), we applied these prediction models to GWAS to assess the association between genetically predicted alternative splicing and cancer risk.

### Annotation of the identified genes using cancer-related gene database

To verify the evidence whether the TWAS-identified genes are related to cancer susceptibility, we extracted cancer related gene sets from the MGB database. Putative cancer related genes were characterized based on their annotation with the key words ‘breast cancer’, ‘prostate cancer’ and ‘lung cancer’. We calculated the number and percentage (success rate) of putative cancer related genes that overlapped with those extracted from the MGB database among the identified genes in this study. Previous TWAS or eQTL studies for breast cancer (19,28,32,38,53) , prostate cancer (31,38,56–58) and lung cancer (38,58,59) reported genes related with these cancers. Genetic variants related with risk of breast cancer (54,55), prostate cancer (99) and lung cancer (85,100) were reported in previous GWAS. We also examined the overlapping between the genes identified in this study with predisposition genes, cancer driver genes and CGC-based gene sets. To evaluate whether our identified genes significantly enriched in these cancer-related genes, we conducted enrichment analysis based on the probability mass function of the hypergeometric distribution. Similar to our previous work (38), the *P*-value is calculated as phyper function implemented in R.

## Data Availability

Supplementary Data 1 provides the download information for the summary statistics of GWAS data for breast cancer, prostate cancer, lung cancer, and three brain disorders (SCZ, ASD, and AD); the epigenetic data, including ChIP-seq data of transcription factors, DHSs, enhancer, promoter, 3D genomics informed regions, enhancer gene links, and eQTLs used in this study; and The functional annotation data, including target cancer related genes, CGC, and cancer driven genes. Gene expression and alternative splicing data generated in breast, prostate, lung and brain tissues, were downloaded from GTEx consortium, and the individual-level genotype was downloaded from dbGaP (https://www.ncbi.nlm.nih.gov/projects/gap/cgi-bin/study.cgi?study_id=phs000424.v8.p2). Gencode annotation (v26.GRCh38) was downloaded from https://www.gencodegenes.org/human/release_26.html. The data from the 1000 Genomes Project data was downloaded through the website, https://www.genome.gov/27528684/1000-genomes-project. For data of essentiality for proliferation and survival of cancer cells, we downloaded wo comprehensive datasets including “sample_info.csv” and “Achilles_gene_effect.csv” from the DepMap portal (Supplementary Data1). Remaining data sources and results are provided within the Article or Supplementary Tables.

## Code Availability

The developed pipeline and main source R codes used in this work are available from Github website: https://github.com/theLongLab/transTF-TWAS or https://github.com/XingyiGuo/transTF-TWAS/.

## Data Availability

Supplementary Data 1 provides the download information for the summary statistics of GWAS data for breast cancer, prostate cancer, lung cancer, and three brain disorders (SCZ, ASD, and AD); the epigenetic data, including ChIP-seq data of transcription factors, DHSs, enhancer, promoter, 3D genomics informed regions, enhancer gene links, and eQTLs used in this study; and The functional annotation data, including target cancer related genes, CGC, and cancer driven genes. Gene expression and alternative splicing data generated in breast, prostate, lung and brain tissues, were downloaded from GTEx consortium, and the individual-level genotype was downloaded from dbGaP (https://www.ncbi.nlm.nih.gov/projects/gap/cgi-bin/study.cgi?study_id=phs000424.v8.p2). Gencode annotation (v26.GRCh38) was downloaded from https://www.gencodegenes.org/human/release_26.html. The data from the 1000 Genomes Project data was downloaded through the website, https://www.genome.gov/27528684/1000-genomes-project. For data of essentiality for proliferation and survival of cancer cells, we downloaded wo comprehensive datasets including sample_info.csv and Achilles_gene_effect.csv from the DepMap portal (Supplementary Data1). Remaining data sources and results are provided within the Article or Supplementary Tables.

## Acknowledgments

We thank GTEx, TCGA, ENCODE, Roadmap, BCAC and other GWAS consortia for providing valuable data resources for this study. The data analyses were conducted using the Advanced Computing Center for Research and Education (ACCRE) at Vanderbilt University.

## Author contribution

X.G. and Q.L. conceived and designed the study. J.H., Q.L. and X.G. performed data collection and processing, bioinformatics and statistical analyses, with additional data preparation and discussion from W.W., J.P., and C.Z.. J.H., Q.L. and X.G. wrote the manuscript with contributions from all other authors. All authors have reviewed and approved the content of the article.

